# PCR test positive rate revealing the real infection epidemic status examined in the COVID-19 epidemic in Tokyo, Japan

**DOI:** 10.1101/2021.09.15.21263609

**Authors:** Toshihisa Tomie

## Abstract

In order to come up with effective infectious disease countermeasures, we need a method to evaluate the effectiveness of the countermeasures. As the most reliable epidemic diagnosis, we propose the PCR test positive rate. Examining the COVID-19 epidemic in Tokyo shows that the positive rate follows the epidemic accurately, without being affected by medical institution closures or the psychology of people. By comparing with the change of the positive rate, we argue that the psychology of people was the cause of the observed asymmetry and shallow valleys in the number of confirmed cases. We discuss that the positive rate as large as 1-5% in the valleys of the epidemic implies that many people are already infected and immune. The observed fact that the positive rate followed the number of positives accurately and the value lower than those reported in the test-negative design studies of influenza are attributable to the government’s policy of finding as many patients as possible. We mention three possibilities as the causes of the repeated COVID-19 epidemic in Japan.

## 1. Introduction

People’s actions were restricted by such as lockdowns of cities to control COVID-19 infection. To evaluate the effectiveness of countermeasures, we need to understand the status of the infection epidemic correctly and promptly. It is common to analyze the number of the PCR (polymerase chain reaction) test positives. The first COVID-19 epidemic in Wuhan ended as forecasted by analyzing the change in the number of positives (ref.1). However, there are some problems in the use of PCR test positives.

The major problem is that we cannot wipe out the doubts about whether the number of PCR test positives correctly reflects the epidemic. It is not easy to find all potentially infected people including asymptomatic people, but it is not necessary. To estimate the effectiveness of the disease control, it is sufficient for any index to be in proportion to the epidemic. However, it is not easy to verify whether the index reflects the real epidemic. When forecasting the end of the ever-first COVID-19 epidemic in Wuhan, we found that the number of infected people announced was initially lower than the forecast curve (ref.2). Our speculation is the following. Initially, people did not pay much attention to the disease, resulting in fewer PCR tests and not finding certain infected people. On the other hand, when people hear the news that the number of confirmed cases is increasing sharply, people might become anxious about their infection, and many people may take PCR tests. An increase in the PCR tests increases the number of positives, making the number of positives un-proportional to the epidemic. Thus, the number of PCR test positives might be affected by the psychology of people.

The number of positives also fluctuates due to the non-constant testing capacity of medical and testing institutions and the processing capacity of test result processing institutions. On holidays and public holidays, the number of inspections reduces significantly. More people will visit hospitals before and after holidays, and the number of positives can be larger before and after holidays. For reducing this fluctuation, it is common to perform a 7-day moving average, but a long-term moving average impairs the time resolution of the information of the epidemic and hinders the correct evaluation of the effect of policy.

The author devised a day of the week correction method as a smoothing method that does not impair the time resolution. If the number of tests reduces every Sunday, for example, to a half, the fluctuation can be smoothed out by doubling the number. This method provides a smooth epidemic curve without the time resolution being impaired. However, it is not easy to compensate for the effects of holidays other than Sundays.

The number of deaths can be used to solve the above problems (ref.3). There is no missing the number. In addition, the number of deaths is not affected by the closures of hospitals or the psychology of people. However, under the age of 70, the disease mortality decreases exponentially with age. According to the data, the mortality rate of COVID-19 is 25% in the 90s and over, 19% in the 80s, 7.5% in the 70s, 2.2% in the 60s, 0.51% in the 50s, 0.2% in the 40s, and 0.03% in the 30s (ref.4). Therefore, analysis of the death number can find older people with higher mortality rates, but not younger infected people with lower mortality rates. The percentage of infected people over the age of 70 is less than 10%.

More seriously, it takes a long time from infection to death. Reference 3 reported that the change in the number of deaths was 23 days behind that of the confirmed cases. Reference 5 reported that the change in the number of infected people delayed about two weeks behind the actual infection. Therefore, the change in the number of deaths is information that is one and a half months behind the actual infection. In some cases, the epidemic ends before that. Therefore, analyzing the number of deaths does not help the timely evaluation of infectious disease countermeasures.

We propose the PCR positive rate as the most reliable epidemic diagnosis for understanding the epidemic status by solving the above problems. The positive rate is a ratio of the positives to the number of the PCR tests. Examining the COVID-19 epidemic in Tokyo shows that the positive rate follows the epidemic accurately without being affected by medical institution closures or the psychology of people.

By comparing with the change in the positive rate, we discuss that the asymmetry and the shallow valleys in the number of confirmed cases were caused by people’s psychology. From the fact that the positive rate did not go below 1-5% in the valleys of the epidemic, we discuss that many people are already infected and immune. The reasons for the positive rate following the number of positives accurately and for the observed fact that the value at the peak of the epidemic was lower than those reported in the test-negative design studies of influenza are attributed to the government’s policy of finding as many patients as possible. Three possibilities are mentioned as the causes of the repeated COVID-19 epidemic in Japan; 1. a short lifetime of immunological memory cells, 2. quick mutation of the virus, 3. people getting sick due to sudden changes in climate show symptoms.

## 2. Method

All data used for analysis in this paper are publicly available. In both Tokyo and Osaka, data on the daily number of PCR tests and positives were obtained from the “Coronavirus Infectious Disease Control Site” (ref.6, ref.7). Israeli data were obtained from the Israeli Ministry of Health website (ref. 8). Weekly patients of various infectious diseases of the country, weekly patients of influenza in Tokyo, and the daily numbers of CVID-19 confirmed cases and deaths were obtained from refs.9, 10, and 11, respectively.

## 3. Results

### 3-1. day-of-week correction of confirmed cases for eliminating weekly dips

Figure 1(a) shows the changes in the PCR test positive rate and the number of positives in Tokyo since March last year (ref.6). The positive rate is displayed linearly, and the number of positives is displayed in a log scale. Figure 1 (b) shows an enlarged view of this summer’s change. In the curve of the number of positives shown by black squares, there are deep dents with one week period, reflecting the decrease in the number of tests on Saturdays and Sundays, but the fluctuation in the positive rate shown by red squares is small. By multiplying the number of positives detected on Sunday by 2.3, Saturday by 1.4, and Monday by 1.2, the deep dents of one week period in the number of positives seen in Fig. 1 (b) can be eliminated. The positive rate on Sunday was multiplied by 0.75 to reduce the weekly spikes in the curve of the positive rate. Figure 1 (c) shows the results of the day-of-week correction. There is a deep dent on July 22nd and 23rd in the change in the corrected number of positives. This dip is considered to be due to the special four consecutive holidays, in which two holidays were moved to coincide with the opening ceremony of the Tokyo 2020 Olympics. The big dent on August 9th will be due to summer vacation. These structures are not seen in the positive rate.

**FIG.1:**
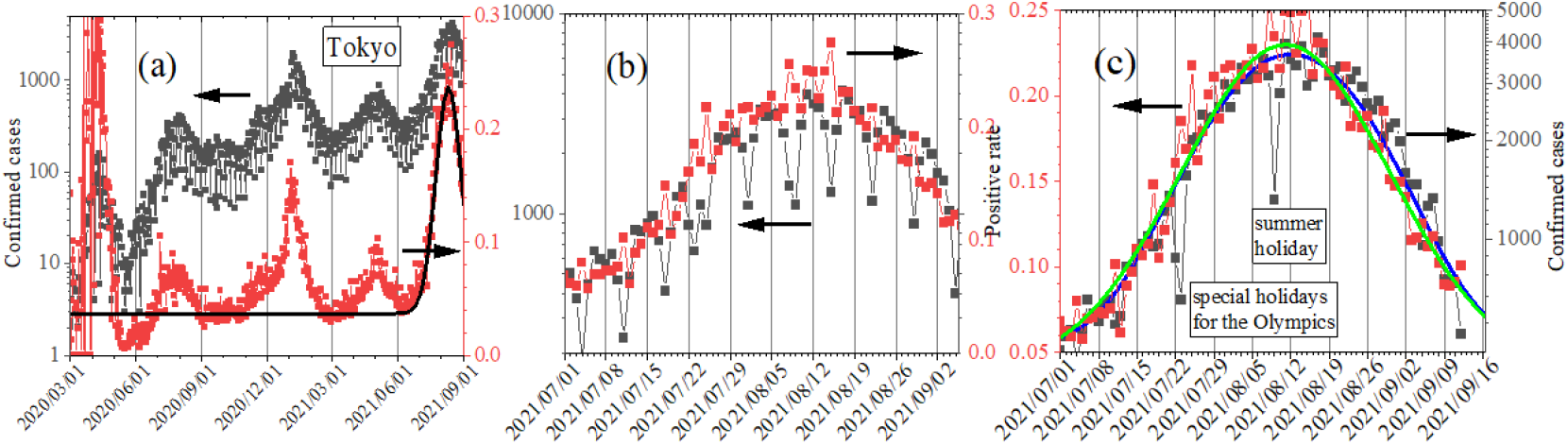
(a) The whole history of confirmed cases (black squares) of COVID-19 and positive rate (red squares) in Tokyo (ref.6). (b) There are deep weekly dips in the confirmed cases (black squares). (c) The day-of-week correction removes weekly deep dents in the change of the number of the confirmed cases.

As shown in Fig. 1 (c), the change in the positive rate exactly agrees with the change in the number of positives. Thus, we confirm that the positive rate gives a real epidemic of infectious diseases not affected by the decrease in the number of tests due to hospitals’ closures.

The change in the number of positives is fitted with a Gaussian of

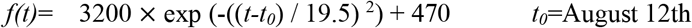

and the change in the positive rate is fitted with a Gaussian of

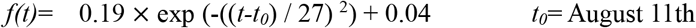

Both changes have a constant value of 470 people for the confirmed cases and 4 % for the positive rate.

As shown in Fig.1 (c), the August epidemic began in July, slowed in August, and peaked on August 11. Assuming that it takes two weeks from the actual infection to the PCR test results (ref. 5), the infection actually started in mid-June, slowed in mid-July, and peaked at the end of July. As mentioned above, the Tokyo Olympics started on July 23 and ended on August 8. One claim is that the Olympics have curbed the epidemic. However, this argument is not acceptable, as this epidemic is well fitted by the Gaussian theoretically expected for natural infections.

### 3-2. change in the number of deaths in the epidemic

Figure 2(a) shows the changes in the number of positives and deaths in Tokyo (ref.11). With the number of deaths multiplied by 50 and with shifting the horizontal axis to the left by 30 days, both changes in the number of positives and deaths agree quite well, meaning fatality is 2 % and that people die 30 days after infection on the average. The population of Tokyo is 14 million, which is comparable to those of small countries; a little smaller than the Netherland, but larger than Sweden, and two to three times those of Denmark and Ireland. Still, the number of deaths is small, fluctuates largely, and reliable analysis is difficult. Figure 2(b) shows the changes in the number of positives and deaths for all of Japan (ref.11). The horizontal axis of the change in the number of deaths was shifted to the left by 25 days. Now the population is as large as 126 million and the fluctuation of the number of deaths is acceptable for some analysis. Until June 2021, the number of PCR test positives was 50 times the number of deaths, but from July the number of positives increased to 200 times the number of deaths. The sudden drop of the fatality to one quarter was caused by a change in the age distribution of infected individuals. The percentage of positives aged 70 and over, who have a high fatality was 8% in May and 5% in June but decreased to 2% in July and 3% in August (ref.12).

**FIG.2:**
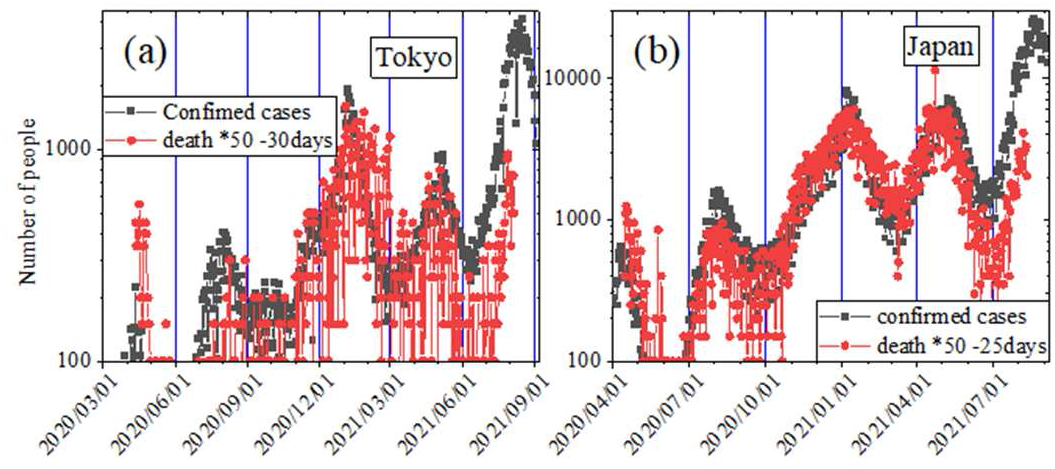
Confirmed cases (black squares) and deaths (red squares) of COVID-19 in (a) Tokyo and (b) over Japan (ref.11). The number of deaths is multiplied by 50 for Tokyo, and by 200 for Japan. Deaths delayed by 30 days for Tokyo and 25 days for Japan from the confirmed cases.

Because the number of deaths is so small, the fluctuation of the data is large and the number of deaths is not suitable for detailed analysis of an epidemic. More seriously, the information is one month older than the change in the positives, so it is nearly useless for a quick evaluation of infectious disease countermeasures.

### 3-3. effect of Golden Week in May

Figure 3 (a) shows the weekly reports of the most prevalent infectious diseases in Japan (ref. 9). There are two deep dents in the 1st and 18th weeks. In Japan, there are multiple holidays between the end of April and the beginning of May in about 10 days. This period is called the Golden Week, and some people take long vacations. Hospitals are often closed. The dent at the 18th week is due to the Golden Week. The effect of the Golden Week is also noticed in this year’s COVID-19 epidemic in Tokyo.

**FIG.3:**
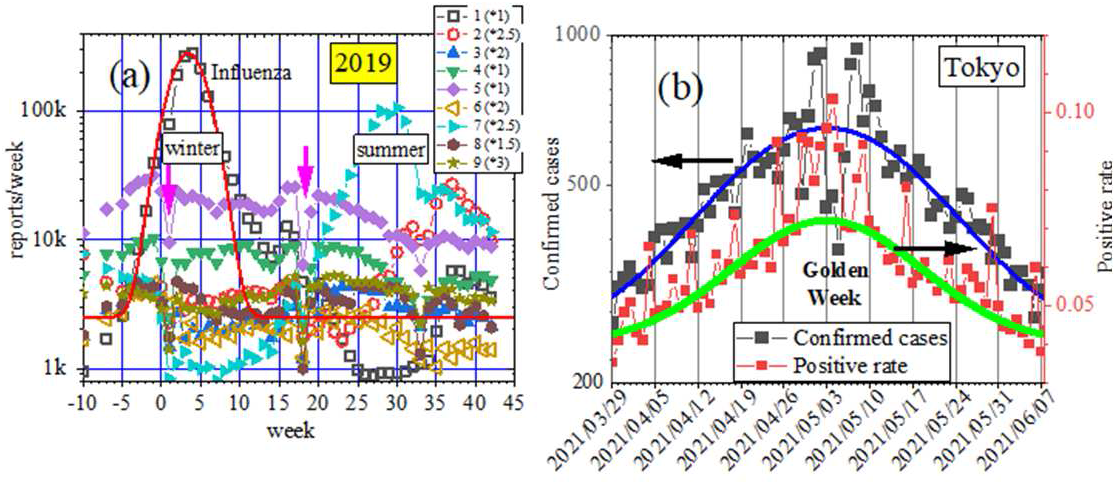
(a) Weekly reports of the most prevalent infectious diseases in Japan (ref.9); 1: Influenza, 2: RS virus, 3: Pharyngoconjunctival fever, 4: Streptococcal pharyngitis, 5: Infectious gastroenteritis, 6: Water sputum, 7: Hand-foot-and-mouth disease, 8: Infectious erythema, and 9: Idiopathic exanthema subitem. There are deep dents in the 1st week and 18th weeks. (b) The deep dent in the 18th week due to the Golden Week in the change of the day-of-week corrected confirmed cases of COVID-19 (black squares) in Tokyo is not seen in the change in the PCR positive rate (red squares).

The black squares in Fig. 3(a) show the day-of-week corrected number of positives. There is a deep dent between May 3rd to 6th due to the closure of hospitals. On the other hand, there is no dent of the Golden Week in the positive rate shown by the red squares. A peak is seen in the change in the positive rate from April 29th to May 6th. Because the duration of the peak was only one week, it may not be a real epidemic. Because people who got inspected during the Golden Week might have slightly heavier symptoms, the PCR test positive rate of these people may be high.

### 3-4. effect of New Year holidays

In Japan, December 29th to January 3rd is the New Year holidays. People return to their hometowns and meet relatives. During this period, all schools are closed and many companies and most hospitals are also closed. The dip at the 1st week in Fig. 3 (a) corresponds to the New Year holidays.

Figure 4(a) shows the change in the number of influenza patients in the 2014/15 season in Tokyo (ref.10). The main patients with influenza are 5 to 9 years old children. The number of influenza patients in the New Year holidays was about one-fifth of that in other weeks.

**FIG.4:**
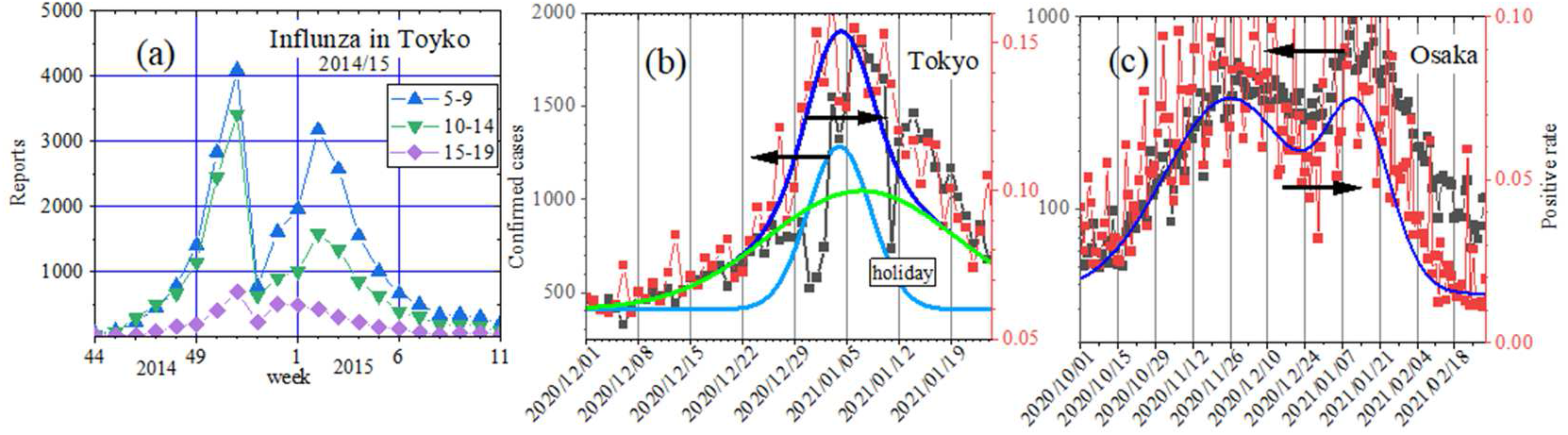
(a) Weekly reports of influenza in the 2014/15 season in Tokyo (ref.10). (b) The day-of-week correction removes weekly deep dents in the change of the number of the confirmed cases. (c) Positive rate and day-of-week corrected confirmed cases of COVID-19 in Osaka(ref.7).

The impact of the New Year holidays was great in the number of PCR test positives of the COVID-19 epidemic. The number of confirmed cases increased sharply on January 5th all over the country. In the case of Tokyo, the number was 600 people on January 3rd and was 1900 on 6th. Figure 4(b) shows the change in the day-of-week corrected number of confirmed cases in Tokyo. We find a drastic decrease in inspections on December 31st. A deep dip on January 11th corresponds to a national holiday. In the change in the positive rate shown by the red squares, there is no dent caused by the holidays.

This summer (Fig. 1(c)) and spring (Fig. 3(b)) epidemics are fitted with a single Gaussian, but two Gaussian were needed to reproduce the winter epidemic (Fig. 4(b)). One epidemic continued for two months starting from the beginning of December and ending at the end of January with a time constant of 18 days, and the duration of the second epidemic was 6 days from the end of December to the beginning of January. These epidemics cannot be identified in the change in the number of positives because the influence of the New Year holidays was very large. As shown in Fig. 4(c), there are two Gaussians also in the positive rate in Osaka (ref.7). The epidemic with the peak in January had a time constant of 17 days and the epidemic with the peak time at the end of November had a time constant of 35 days. Not only in Tokyo and Osaka, but throughout Japan, the winter epidemic from late October to February was highly regional.

### 3-5. special characteristics of the first CVID-19 epidemic in Spring 2020

The first epidemic in Japan last spring differs from subsequent epidemics in several points. First, as shown in Figure 5(a), the peak of the change in the number of positives was one week later than the peak of the positive rate in Tokyo. As seen in Figure 5(b), the peak date of the change in the number of positives was 6 days behind the peak of the positive rate in Osaka. In subsequent epidemics, as seen in Figures 1(c), 3(b), 4(b), and 4(c), the number of positives and the positive rate peaked on almost the same day.

**FIG.5:**
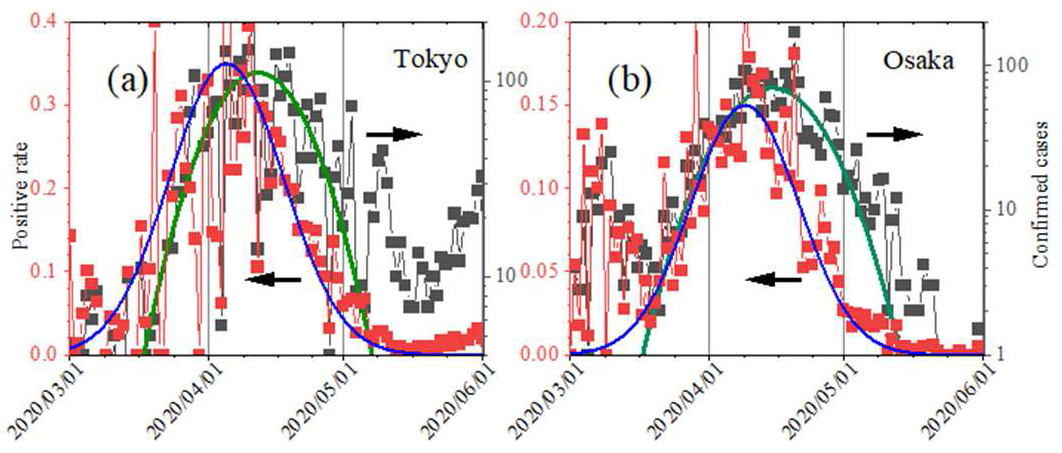
In the first epidemic, the peak of the number of the confirmed cases (black square) was delayed by one week from the peak date of the positive rate (red square) and the changing profile was asymmetry both in (a) Tokyo and(b) Osaka.

Second, the change in the number of positives was asymmetric while the change in the positive rate was symmetric. In other epidemics as seen in Figures 1(c), 3(b), 4(b), and 4(c), the change in the number of positives can be fitted with a Gaussian, but only in the first epidemic seen in Fig. 5, confirmed cases decayed slower than the Gaussian shown by the green curve. In Osaka, the asymmetry was a little smaller but the number of confirmed cases was larger than the Gaussian in May.

In detail, there is also a small asymmetry in this summer’s epidemic. In Figure 1(c), the number of positives in late August is slightly higher than that of the Gaussian, which is discussed later in the 4. Discussions

Third, the peak value of the positive rate was high. The peak value of the positive rate was less than 10% in last summer and this spring epidemics, 15% in this winter (Fig. 4(b)), and 22% this summer (Fig. 1(c)), but in the first epidemic shown in Figure 5(a), the peak value of the positive rate was around 35%. In Osaka, the peak value of the positive rate in the first epidemic was 15% (Fig. 5(b)).

### 3-6. the number of confirmed cases and the positive rate in the valleys of epidemic

As seen in Fig. 1(a), there are five peaks in spring, summer in last year, winter, spring, and summer this year. In May last year after the first epidemic, the number of positives decreased to about 10 as seen in Fig. 5(a), but after that, in the valleys of the second and later epidemic, confirmed cases were as large as 150, 200, and 250 in October, March, and June, respectively. The number of positives in the valley of the second and subsequent epidemics was comparable to or higher than the number of positives in the peak of the first epidemic. As shown in Fig. 6, the same thing happened in Osaka (ref.7).

**FIG.6:**
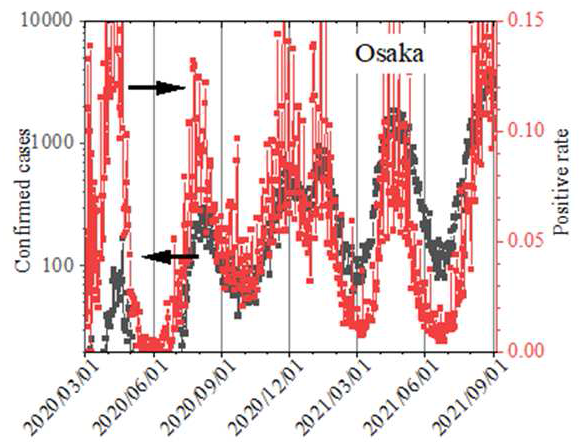
The whole history of the confirmed cases (black square) and positive rate (red square) in Osaka(ref.7). The number of confirmed cases in the valleys was high except after the first epidemic.

Similarly, the positive rate decreased to almost zero in May after the first epidemic as seen in Fig. 5(a), but in the valleys of the second and later epidemic in October last year and March and June this year the positive rate decreased only to 4%. Also in Osaka, the positive rate decreased to 0% after the first epidemic as seen in Fig. 5(b), but after that, the positive rate in the valleys of the epidemic was 1% to 3% as seen in Fig. 6.

The different behavior of the positive rate from the confirmed cases is that the values in the valleys are well smaller than the peak value in the first epidemic.

## 4. Discussions

### 4.1 epidemic of a single pathogen in a single community

When the population and microorganisms increase exponentially at the beginning, the rate of increase in the number of individuals decreases and converges to a steady value as the number approaches the capacity of the environment, and this phenomenon is described by the logistic equation (ref.16).

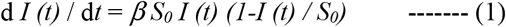

Here, *I (t)* is a cumulative value of the increased number of individuals at time *t, S*_*0*_ is the saturation value, and *β* is the rate of increase.

In the case of infectious disease transmission, it is no longer the source of infection once the infected person recovers or is quarantined. Kermack and McKendrick (ref.17) made a modification incorporating the effect.

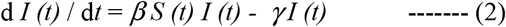

Here, *γ* is the rate at which an infected person is quarantined or recovered, and *S (t)* is the number of uninfected persons at time *t*, given by the following equation.

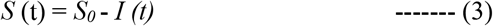

Both the logistic function and numerical solutions of K-M equations can be approximated by a Gaussian (ref.2).

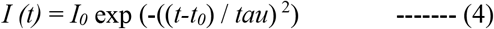

Therefore, the epidemic of infectious diseases should be nearly symmetrical and approximated by a Gaussian. The above theory applies to an epidemic of a single source of a pathogen in a single community. When multiple communities have epidemics of different parameters; different peak dates, *t*_*0*_, and different time constants, *tau*, in Eq. (4), the total epidemic is a sum of all epidemics. In fact, our success in forecasting the end of the first epidemic in Wuhan (ref.1) owed to the separation of epidemics in three regions; Wuhan city, Hunan province except for Wuhan, and the whole of China except for Hunan province. When there are plural sources of the pathogen and when they are not separated, the total epidemic is described by a sum of Gaussians describing the epidemics of individual sources. In some cases, there may not appear a sharp peak in the total epidemic.

### 4.2 the asymmetry in the change in the number of confirmed cases caused by psychological factors

When an epidemic cannot be described by a single Gaussian, we have to consider fitting the epidemic by plural Gaussians as we did for forecasting the end of the epidemic in Wuhan (ref.1). However, if the change in the number of patients is different from the change in the positive rate, the cause of the deviation of the change in the number of patients from a Gaussian should be a result of some non-infectious effects.

The positive rate of this summer’s epidemic in Tokyo in Fig. 1(c) can be fitted with a Gaussian. Therefore, we could understand that this epidemic was caused by a single source of a pathogen. However, the change in the number of positives deviated from the Gaussian after the peak. To make it easier to see, Fig. 7 shows an enlarged view of the change after the peak. The epidemic was peaked on August 12. The confirmed cases started to deviate upward from the Gaussian from August 19, one week after the peak and returned to the Gaussian on September 3. The number of positives was larger than the Gaussian only for two weeks. The deviated area is shown in orange.

**FIG.7:**
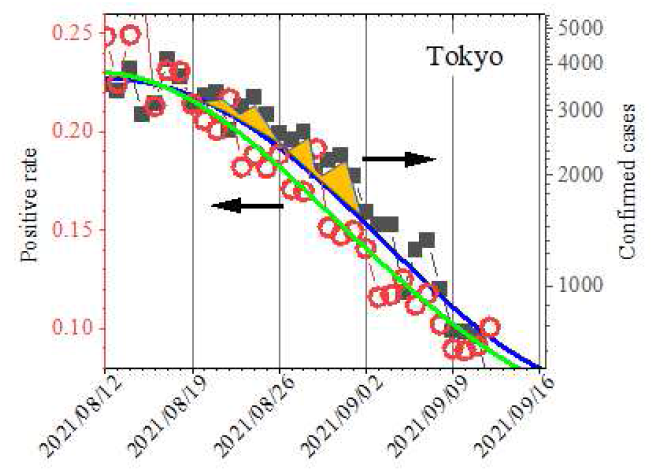
An enlarged display of Fig.1 (c) after the peak of the summer epidemic. The number of positives deviated upward from the fitting Gaussian (orange part) for two weeks from Sep. 19 one week after the peak.

The increase in positives above the Gaussian can be due to psychological effects. Immediately after the peak, many were surprised by the surge in infected people, worried about themselves, and had their PCR tests. When the number of confirmed cases decreased to one-third of the peak, people were relieved and the number of PCR tests returned to normal. Of course, the number of positives was not false in this case as well. But the change in the number did not reflect the real epidemic. We get information on an epidemic from the shape of the change, and the shape of the change should be determined only by the epidemic, not by psychology. The percentage of people taking PCR tests can be any number but the essentially important thing is that the percentage should be always constant for knowing the real status of an epidemic.

A psychological effect was also observed in the confirmed cases in the first epidemic last year April in Tokyo (Fig. 5(a)). The change in the number of positives was asymmetry. The number of positives increased 10 times in about three weeks and peaked in mid-April. But it took five weeks for the number of positives to decrease to one-tenth of the peak value. This asymmetry can be explained as in the case of August this year discussed above. Asymmetry was noticed also in the first epidemic in Osaka. The number of the confirmed cases was larger than the Gaussian shown by the blue curve in May 2020 as seen in Fig. 5(b).

In the first epidemic last spring, the peak number of positives was one week behind the peak positive rate, with marked asymmetry as shown in Figure 5. On the other hand, the asymmetry of this summer’s epidemic seen in Figure 7 is small and can only be recognized by detailed analysis. This may mean that people were very afraid during the first epidemic, but that people are more reassured after experiencing some epidemics.

The asymmetrical change in the number of infected people observed in Israel could be also attributable to a psychological effect. Figure 8 shows changes in the number of infected people and the positive rate in Israel (ref. 8). The number of positives (black squares) is displayed in a log-scale with the range from 400 to 4000 so that it changes in the same range as the positive rate (red squares) displayed linearly from December last year to March this year. The changes in the number of positives and the positive rate were in perfect agreement. The change in the positives of the epidemic in January 2021 is fitted with a Gaussian of

**FIG.8:**
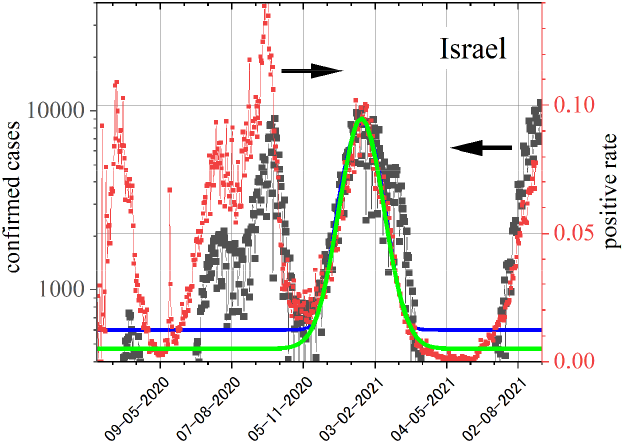
The whole history of the confirmed cases (black square) and the positive rate (red square) in Israel (ref.8). Quite a small number of positives in the June valley can be attributable to people’s feeling of safety by the progress of vaccination.

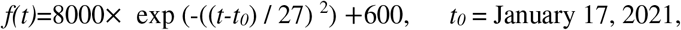

and the change in the positive rate is fitted with a Gaussian of

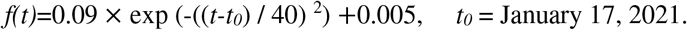

However, the number of positives increased greatly over the Gaussian in March after the peak, which can be a psychological effect.

### 4.3 absolute value of the positive rate

We know the positive rate generally expected in PCR tests for infectious diseases from researches on the test-negative design applied to influenza.

The test-negative design is one of the methods to evaluate the effect of some measures for a certain disease. The PCR test is performed against patients who visit a medical institution with showing symptoms similar to the symptoms of the disease of interest. The effect of some measures is evaluated from the number of positive and negative patients. This method has become a standard method for evaluating the infection control effect of influenza vaccines.

In one research, an infection suppression rate of 36% was concluded for the 2017/18 season in five states of the United States (ref. 13). Among 4562 patients who visited medical institutions for influenza-like illness, 2259 were vaccinated. Among vaccinated people, 741 were PCR positive. Then, the positive rate was 33%. Among 2303 non-vaccinated people, 97 were PCR positive. Then, the positive rate was 42%.

In another research for the 2015/16 season, an infection suppression rate of 48% was concluded (ref. 14). Among 3396 vaccinated people, 492 were PCR positive, and among 3483 non-vaccinated individuals, 817 were PCR positive. Therefore, the positive rates were 14.5% and 23%, respectively.

In Japan, there is a report of the test-negative design for children under 6 years old (ref.15). Positive/negative = positive rate in 4 seasons of 2013/14, 2014/15, 2015/16, and 2016/17 were 386/435 = 0.47, 303/555 = 0.35, 424/490 = 0.46, and 369/640 = 0.37, respectively.

From these researches, we know the PCR positive rate will be from 30% to 50% when patients suspected of a certain disease are PCR tested.

From the values reported in the above papers, we understand the positive rate of the first COVID-19 epidemic in Tokyo was reasonable. However, the positive rate of 6% last summer, 15% at the beginning of this year, and 10% this summer are quite small. This is considered to reflect the policy of the Japanese government. In the researches mentioned above, the test-negative design for influenza targets people who develop influenza-like symptoms. However, the COVID-19 PCR tests are performed against the close contacts of already found positives in order to find as many infected individuals as possible. The positive rate is inevitably low because the test is done even if the patient is asymptomatic. The less likely an infected person is overlooked, the lower the positive rate.

In Israel shown in Fig.8, the positive rate at the peak of the epidemic last September was 14%, and the positive rate at the peak of other epidemics was 10%. Because the differences of the positive rate at the peaks and in the valleys in Tokyo of about 6% is very small compared to Israel, we can say that Japan’s attitude to COVID-19 is more nervous than Israel.

While the positive rate was over 30% in the first epidemic in Tokyo, it was about 10% in Israel. This might indicate that last spring the Israeli government was more cautious than Japan.

### 4.4 the positive rate and the number of infected people in the valleys of the epidemic

After the first epidemic last spring, the positive rate decreased to almost zero % in Tokyo (Fig. 5(a)), Osaka (Fig. 5(b)), and Israel (Fig. 8). However, after the second epidemic and later, the positive rate in the valleys remained as large as 1 to 4%. Since the influence of psychological factors on the positive rate is negligible, the non-zero positive rate in the valleys implies that there are always coronavirus carriers among people even when COVID-19 is not in an epidemic.

Some reports support our hypothesis. For eight days until April 7 in 2020, the research team at Kobe City Medical Center General Hospital conducted a blood test on 1,000 outpatients who visited the outpatient department, and 3.3% of them had antibodies (ref.18). From April 13 to 19 in 2020, Keio University Hospital (Tokyo) conducted PCR tests on 67 patients who were hospitalized for treatment of diseases other than COVID-19 and had no symptoms such as fever and four were positive, and the positive rate for PCR test of COVID-19 was 6% (ref.19). In Okinawa, a mass test of 2064 restaurant employees was conducted on August 1st and 2nd 2020, and 86 were positive. The positive rate was 4% (ref.20).

In the case of Okinawa, August when the test was conducted was after the first epidemic, so we can explain that the epidemic spread the virus among people. April 3rd to 7th when the tests were conducted at the hospital in Kobe was about a week before the peak of the number of positives, April 12th. But the peak date of the positive rate was April 5th. Then, the test in Kobe was performed only a few days before or at the peak of the epidemic. Therefore, it cannot be excluded that the epidemic spread the virus when Kobe hospital conducted the test for antibodies.

In Tokyo (Fig. 5(a)), Osaka (Fig. 5(b)), and Israel (Fig. 8), the number of infected people was almost zero after the first epidemic last spring. However, in the subsequent epidemics, the number of positive people was large even in the valleys. Not like the positive rate, the high values of the confirmed cases in the valleys could be a psychological effect. One reason for doubting the psychological effect as the cause of many positives in the valleys of the epidemic is that the number of the confirmed cases in Israel decreased to as small as 20 in the June valley, while the number was about 700 in the valley after the epidemic in September 2020 (Fig.8). The 20 people in June this year were only one over 500 of the peak value in January just before. The depth of the three valleys in Tokyo last October, March, and June this year is only one-half to one-fifth of the peeks. In Israel, the double vaccination rate of the COVID-19 vaccine exceeded 50% in mid-March (ref.8). People’s sense of security due to the progress of vaccination may have created a deep valley of confirmed cases in the June valley.

In Israel, the positive rate was almost zero from April to June (Fig. 8). This can be the effect of the vaccine. However, to reach a conclusion, the time history of increased vaccination rates and decreased PCR positive rates needs to be analyzed in detail.

### 4.5 considerations on two questions

Finally, we consider two important questions.

#### Question 1: Why does the positive rate increase with the number of patients?

If a doctor decides that a patient with symptoms similar to those of the target disease should undergo a PCR test, the positive rate will be almost 100% if the doctor is competent. Even if the doctor’s ability is not very high, the positive rate will be 30% to 50%. In any case, the positive rate is almost constant regardless of the epidemic.

Our hypothesis to explain the observed fact that changes in the positive rate of COVID-19 were in perfect agreement with changes in the number of confirmed cases is as follows.

PCR tests to find COVID-19 patients are performed almost randomly, not just in patients with COVID-19-specific symptoms. If the test is random, the higher the number of patients in the epidemic, the more likely it is to find a patient, leading to a higher positive rate.

#### Question 2: Why is the epidemic repeated?

In the K-M equation (ref.17) shown in Eq. 2, the rate of increase in the number of infected people is given by *(β S (t) - γ) I (t)*. When the number of infected people increases and the number of uninfected persons, *S (t)* = *S*_*0*_*-I (t)*, decreases, the product of the number of uninfected persons and the rate of increase *β* becomes smaller than the rate of decay *γ, β S (t) - γ <0*, and then, the epidemic begins to end. The epidemic can resume only when the infected person, *I (t)*, turns into an uninfected person, *S (t)*.

Antibodies are produced in the body of infected persons and prevent the virus from entering again. Although the lifetime of antibodies is not so long, immunity is maintained for a long time when long-lived immunological memory cells are produced (refs.21 - 24). Therefore, the epidemic will not repeat as long as the memory cells do not lose their ability.

However, it is not uncommon for immunized infectious diseases to repeat epidemics. Rather, most infections are recurring. Common cold with multiple pathogens is repeated 1.5 times a year in adults (refs.25–28). Most infectious diseases do not have sharp epidemic peaks, as seen in Fig. 3(a). Influenza affecting 10% of Japanese people every year has a sharp peak in winter, but there are non-negligible patients throughout the year.

Infectious diseases develop without the invasion of external pathogens, which was evidenced without any doubt by the fact that Antarctic winterers who were isolated from the outside world for several months also caught colds (refs.29 and 30).

From the above considerations, there are three possibilities as the cause of repeated COVID-19 epidemic; 1. a short lifetime of immunological memory cells, 2. the virus mutates frequently and the effect of the acquired immunity weakens, and 3. people develop the disease like a common cold.

## 5. Summary

Examining the COVID-19 epidemic in Tokyo proved that the PCR test positive rate can be the most reliable diagnosis of epidemics for revealing the real epidemic status. The change in the positive rate closely followed the positives with no structure caused by medical institution closures which generated deep weekly dips in the number of confirmed cases in the Golden week in May, new Year holidays, and other holidays.

An epidemic of infectious diseases of a single source of a pathogen in a single community is described theoretically by the K-M equation, and its numerical solution can be approximated by a Gaussian. In all five epidemics since the first epidemic in Spring last year in Tokyo, the changes in the positive rate were fitted quite well by Gaussians, which may imply all five epidemics can be an epidemic of a single source of a pathogen.

While it was almost 0 % in the first valley after the first epidemic, the positive rate was as large as 1 to 4 % in the three valleys after the second epidemic, which was attributable to the widespread of the coronavirus among people.

The peak values of the positive rate well lower than those reported for the test-negative design studies of influenza were attributed to the government’s policy of finding as many patients as possible. The positive rate followed the positives closely not only in Tokyo but also in Osaka and Israel. This fact was also attributable to the policy.

On the other hand, many asymmetric phenomena not fitted by a single Gaussian were observed in the number of confirmed cases. In all asymmetries, after the epidemic peak, the number of positives was higher than the Gaussian for fitting the rising part of the epidemic. These asymmetries were attributable to the psychology of people.

Three possibilities were mentioned as the cause of the repeated COVID-19 epidemics in Japan; 1. a short lifetime of immunological memory cells, 2. quick mutation of the virus, 3. people getting unwell caused by climate change develop the disease.

## Data Availability

All data used for analysis in this paper are publicly available.

